# SARS-CoV-2 variants of concern, variants of interest and lineage A.27 are on the rise in Côte d’Ivoire

**DOI:** 10.1101/2021.05.06.21256282

**Authors:** Etilé Augustin Anoh, Grit Schubert, Oby Wayoro, Monemo Pacôme, Essia Belarbi, Andreas Sachse, Sébastien Calvignac-Spencer, Fabian Leendertz, Bamourou Diané, Chantal Akoua-Koffi

## Abstract

SARS-CoV-2 variants of concern (VOC) and variants of interest (VOI) are heavily altering the COVID-19 pandemic’s course due to their increased transmissibility, virulence and immune escape abilities. Data on their spread in western sub-Saharan Africa is however still scarce. We therefore sequenced viral genomes from SARS-CoV-2 cases identified in central and northern Côte d’Ivoire between May 2020 and March 2021. We report the introduction of VOC B.1.1.7 as early as mid-January 2021, followed by detection of a single case of VOC B.1.351, as well as VOI B.1.525. Since early 2021 VOC/VOI are gradually dominating the SARS-CoV-2 virus pool in Côte d’Ivoire, as seen in other regions of the world. Intriguingly, we also find that another lineage, A.27, has also been on the rise over the same period. In sum, this study highlights again the extremely rapid local diffusion of VOC, VOI and possibly A.27, and the importance of further reinforcing capacities for genomic surveillance on the African continent.

## Main Text

The ongoing SARS-CoV-2 pandemic is coined by the emergence and spread of viral lineages with increased transmissibility, virulence and immune escape abilities, the variants of concern [VOC; (1, 2)]. Since end-2020, three VOC - B.1.1.7 which originated in the U.K (20I/501Y.V1), B.1.351 (20H/501Y.V2) from South Africa, and P.1 (20J/501Y.V3) from Brazil - have shown sustained regional community transmission followed by quick international dispersal. B.1.1.7 is now on the verge to becoming the dominant lineage worldwide. The ramping up of genomic surveillance on a global scale has also resulted in the identification of other variants presenting mutations potentially associated with altered phenotypes but not yet formally identified as exhibiting altered properties, the variants of interest [VOI; (2)]. As of March 30^th^, WHO has identified 6 VOI associated to local community transmission and some degree of international spread (1).

SARS-CoV-2 genomic data have accumulated at very different paces in different regions of the world and West Africa is among the least covered [see (3, 4) and references therein]. In order to shed light on the evolution and spread of SARS-CoV-2 and its VOC/VOI in this region, we sequenced viral genomes from cases identified in central and northern Côte d’Ivoire between May 2020 and March 2021.

### Rapid lineage turn-over to VOC/VOI in central Côte d’Ivoire

We obtained nucleic acids extracted from naso/oropharyngeal specimens from the national SARS-CoV-2 surveillance program. Those samples had been tested positive for SARS-CoV-2 by real-time PCR. Between May 23^rd^ 2020 and March 9^th^ 2021, 2637 naso/oropharyngeal specimens from COVID-19 suspect cases had been received by the Centre Hospitalier Universitaire (CHU) Bouaké, of which 524 specimens tested positive for SARS-CoV-2. An additional 8 SARS-CoV-2 positive nucleic acids were obtained from a running surveillance study on acute respiratory infections in the Bouaké region and western Côte d’Ivoire [N_tested_ = 828; (5)].

We generated complementary DNA (cDNA) and performed on-site whole genome sequencing at the CHU Bouaké by applying the multiplex PCR approach described in the ARTIC protocol (6) and sequencing the resulting amplicons on a MinION device (Oxford Nanopore Technologies). We also followed the ARTIC protocol for genome consensus sequence generation (7). Genomes were assigned to viral lineages using pangolin [version 2.3.8, April 2^nd^ 2021; (8)]. Lastly, we also performed additional phylogenetic analyses aimed at refining the placement/lineage assignment of 5 genomes initially assigned to lineage A, of which 4 genomes fell into A.27 diversity and where reassigned accordingly (Supplementary Figure 1). A total of 166 SARS-CoV-2 genomes were generated, spanning the period from May 2020 to March 2021.

From May 2020 to December 2020, we detected 20 pango lineages, with lineage A.19 being by far the dominant one, followed by A.18 and B.1 (Figure 1A and B). From January 2021 on, the distribution of viral lineages changed drastically (Figure 1C). We detected 2 VOCs, B.1.1.7 (first detection January 15^th^ 2021) and B.1.351 (a single detection on March 6^th^ 2021), and one VOI, B.1.525 (first detection February 8^th^ 2021). The frequency of VOC/VOI gradually increased from January to March (Figure 1D – F), leading to a rapid replacement of previously circulating lineages. In March 2021, VOC/VOI altogether represented 67% of sequenced genomes (Figure 1F).

**Figure 1.** Distribution of SARS-CoV-2 lineages among 166 genomes sequenced at the Centre Hospitalier Universitaire Bouaké in Côte d’Ivoire. Data were subdivided into sample collection periods: A) May – July 2020, B) August – December 2020, C) January – March 2021, D) January 2021, E) February 2021, F) March 2021. Viral lineage assignment was conducted using pangolin [version 2.3.8, April 2nd 2021; (6)]. The treemap plot was drawn with R, package treemap (15).

### Emergence and rise of lineage A.27

The emergence of VOC/VOI was accompanied by the appearance and increasing frequency of lineage A.27 (29 % of genomes from January to March 2021; Figure 1D - F). As a number of VOC/VOI do, A.27 harbours many lineage defining mutations [n=18; (9)]. A.27 genomes are characterized by a S:614D background and show four coding mutations in spike potentially linked to increased transmissibility or immune escape [S:L18F, S:L452R, S:N501Y and S:H655Y; (10-12)]. This lineage was initially defined following an outbreak in Mayotte, a French overseas territory and island in the Mozambique Channel (13). As of April 1^st^ 2021, a mere 226 A.27 genomes have been published on the GISAID database, primarily from European countries (14). A.27 was first detected in December 2020 in Denmark but the high proportion of those genomes appearing in France/Mayotte (144 genomes) might pinpoint a potential origin or early elevated spread of the variant in this country. While prior to this study only three A.27 genomes had been reported from continental sub-Saharan Africa (two from Nigeria, one from Rwanda), our results show that that this lineage probably dispersed much more broadly on the continent.

### Limitations of the study

The data presented here are geographically limited to central and northern Côte d’Ivoire and might thus only reflect local outbreak patterns. VOC/VOI/A.27 were probably imported via travellers but the very uneven international sampling prevents firmly determining the origins of importation. Potential epidemiological links to southern Côte d’Ivoire, where Abidjan acts as the main country’s hub for international travel, should be investigated to better understand within country spread.

## Conclusions

The data presented here highlight again the extremely rapid local diffusion of VOC, VOI and, possibly, A.27. Monitoring the spread and possibly local emergence of variants provides information that can guide governmental measures towards pandemic control. In the next few months, reinforcing genomic surveillance on the continent will be a major regional and global task.

## Ethics approval

The study adheres to the tenets of the Declaration of Helsinki and national ethical and legislation standards. The CHU Bouaké laboratory is part of the national surveillance programme for SARS-CoV-2 diagnosis in Côte d’Ivoire. Approval by institutional ethics committees for conducting the ANDEMIA surveillance study was given by the Comité National d’Ethique des Sciences de la Vie et de la Santé de Côte d’Ivoire (ethics permit number 105/MSHP/CNER-dk), and by the Ethikkommission - Ethikausschuss am Campus Virchow-Klinikum, Charité, Germany (ethics permit number EA2/230/17)]. The study objectives were explained by trained surveillance officers, and informed written consent for participating in the study was collected from all participants. Written consent was obtained by parents or legal guardians of minors. Personal patient identifiers were not accessible to the authors. Inclusion of SARS-CoV-2 diagnostics into the study was granted by the Comité National d’Ethique des Sciences de la Vie et de la Santé de Côte d’Ivoire (ethics permit number 073-20/MSHP/CNESVS-kp).

## Data Availability

all sequence data are available on GISAID

https://www.gisaid.org/

## Authors’ contributions

All authors made substantial contributions towards the conduct of the study. CAK, EB, FL, GS designed the study. CAK and BD guaranteed study implementation in Côte d’Ivoire. EB, GS and SCS were driving the development of methodologies and analyses. AS, EAA, GS, MP, OW and SCS performed analyses. EAA, GS and SCS drafted the manuscript. All authors revised earlier versions of the manuscript and approved the final version for submission.

## Informed consent

Suspect cases of COVID-19 were tested for SARS-CoV-2 during clinical routine, following national testing guidelines. Patients were aware of the testing and consented to the sampling of respiratory specimens.

## Data availability

All SARS-CoV-2 sequences generated at CHU Bouaké, Côte d’Ivoire, are available on GISAID.

## Acknowledgements

We are grateful to S. Tomczyk, T. Eckmanns, K. Nowak, P. Pitzinger, S. Kribi and R. Wood for their participation in the development and implementation of the broader study idea, and Caroline Röthemeier and Kevin Merkel for technical support (all Robert Koch-Institute). At Côte d’Ivoire, we thank Prof. M. Dosso, director of the Institut Pasteur of Côte d’Ivoire and national coordinator of COVID-19 laboratory activities, for including CHU Bouaké into respective activities and granting permission to sequencing tested specimens. We further wish to acknowledge Dr. F. Bamba-Touré, the regional director of health at the Ministry of Health, as well as Dr. M. Coulibaly at the Institut National d’Hygiène Publique for facilitating the conduct of the study. Lastly, we extend our thanks to the local SARS-CoV-2 surveillance team of Dr. M. Soundélé, Dr. L. Karidioula, Dr. P. Ouattara, Dr. I. Dembélé, N. Wohi and S. Karidiouala for their efforts in implementing the testing of respiratory specimens for SARS-CoV-2 at CHU Bouaké.

We thank the funding body that made this study possible, the German Federal Ministry of Education and Research (BMBF; grant number 01KA1606 and grant number 01KI2047).

