## Supplemental Figure 1 for "SARS-CoV-2 variants of concern, variants of interest and lineage A.27 are on the rise in Côte d’Ivoire"

Supporting information

**Supplementary figure 1.** Maximum likelihood tree of 144 genomes assigned to the recently emerged SARS-CoV-2 lineage A.27. The tree contains 5 genomes from Côte d'Ivoire that were initially assigned to lineage A, while 4 of them fall into the diversity of A.27.

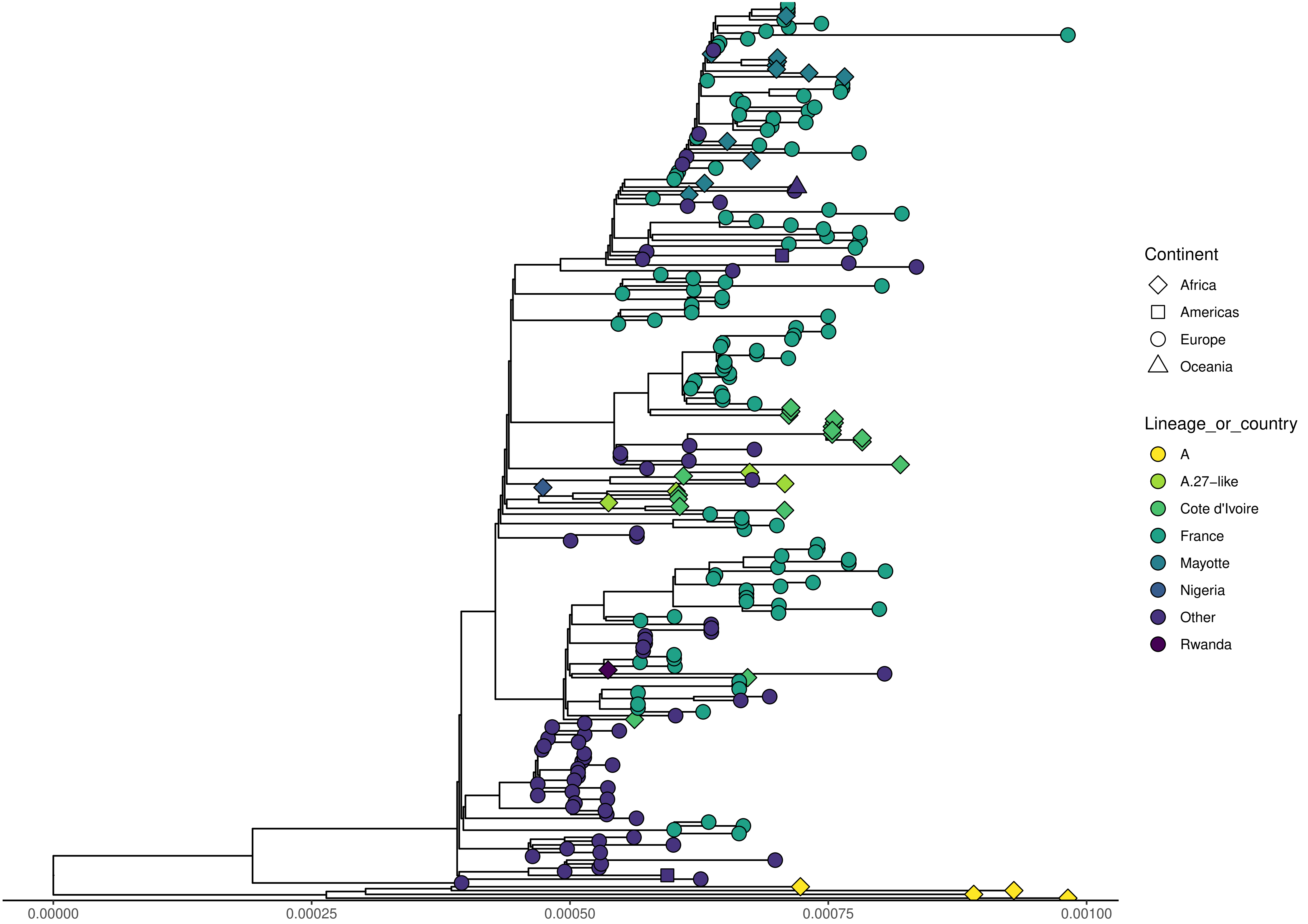
0.00000
0.00025
0.00050
0.00075
0.00100
Continent
Africa
Americas
Europe
Oceania
Lineage_or_country
A
A.27–like
Cote d'Ivoire
France
Mayotte
Nigeria
Other
Rwanda
